# Prospective comparison of Bayesian and frequentist adaptive clinical trials : The SHADOW - SHINE project

**DOI:** 10.1101/2021.06.02.21257838

**Authors:** Kristine Broglio, William J. Meurer, Valerie Durkalski, Qi Pauls, Jason Connor, Donald Berry, Roger J. Lewis, Karen C. Johnston, William G. Barsan, On behalf of the SHINE and ADAPT-IT investigators

## Abstract

**Importance:** Bayesian adaptive trial design has the potential to create more efficient clinical trials. However, one of the barriers to the uptake of Bayesian adaptive designs for confirmatory trials is limited experience with how they may perform compared to a frequentist design.

**Objective:** Compare the performance of a Bayesian and a frequentist adaptive clinical trial design.

**Design:** Prospective observational study comparing two trial designs using individual patient level data from a completed stroke trial, including the timing and order of enrollments and outcome ascertainment. The implemented frequentist design had group sequential boundaries for efficacy and futility interim analyses when 90-days post-randomization was met for 500, 700, 900, and 1,100 patients. The Bayesian alternative utilized predictive probability of trial success to govern early termination for efficacy and futility with a first interim analysis at 500 randomized patients, and subsequent interims after every 100 randomizations.

**Setting:** Multi-center, acute stroke study conducted within a National Institutes of Health neurological emergencies clinical trials network.

**Participants:** Patient level data from 1,151 patients randomized in a clinical trial comparing intensive insulin therapy to standard in acute stroke patients with hyperglycemia.

**Main Outcome(s) and Measure(s):** Sample size at end of study. This was defined as the sample size at which each of the studies stopped accrual of patients.

**Results:** As conducted, the frequentist design passed the futility boundary after 936 participants were randomized. Using the same sequence and timing of randomization and outcome data, the Bayesian alternative crossed the futility boundary about 3 months earlier after 800 participants were randomized.

**Conclusions and Relevance:** Both trial designs stopped for futility prior to reaching the planned maximum sample size. In both cases, the clinical community and patients would benefit from learning the answer to the trial’s primary question earlier. The common feature across the two designs was frequent interim analyses to stop early for efficacy or for futility. Differences between how this was implemented between the two trials resulted in the differences in early stopping.

**Trial Registration:** The SHINE trial was registered and results are reported on clinicaltrials.gov under identifier: NCT01369069

## Introduction

Adaptive clinical trials use interim analyses of the accumulating trial data to make prospectively defined changes.^1^ These changes guide the use of the remaining trial resources in order to most efficiently address the trial’s primary scientific question. Two common adaptive design features in confirmatory trials are sample size re-estimation and early stopping.^1^ These two design features both aim to align the total study size with the number of participants needed to answer the question. Both frequentist and Bayesian approaches to SSR and early stopping have been used in modern confirmatory trials. Additional innovative adaptive design features are increasingly feasible with advances in statistics and clinical trial operations. Bayesian statistical methods have contributed to this recent innovation as they are inherently synthetic, making use of different sources of information, such as both intermediate and final endpoint assessments, and providing intuitive probability statements which can be aligned with scientific priorities.^2^

Despite the potential of Bayesian methods and adaptive clinical trial designs, there has been slow uptake of innovative design features in confirmatory phase trials and also particularly in academic-led research efforts.^3^ To address this, the United States (US) Food and Drug Administration (FDA) and the Office of the Director of the National Institutes of Health (NIH) launched the Advancing Regulatory Science initiative. One project funded under this program was a public-private partnership called the Adaptive Designs Accelerating Promising Trials into Treatments (ADAPT-IT) project.^4^ In ADAPT-IT, academics from an NIH funded neurologic emergency clinical trials network collaborated with other scientists with the objectives of designing Bayesian adaptive confirmatory phase neurologic emergency clinical trials and using mixed methods to gain insights into the process.^5^

The projects from ADAPT-IT are described elsewhere.^6–8^ The investigators planning the Stroke Hyperglycemia Insulin Network Effort (SHINE) trial were the first to participate in ADAPT-IT. A frequentist group sequential trial design with a blinded sample size re-estimation had been developed and funded, and that trial design was retained and executed. However, the ADAPT-IT process resulted in a second, fully specified, alternative adaptive design that was fully Bayesian. The Bayesian adaptive design used more frequent interim analyses to assess the trial for early stopping. Both designs were fully and rigorously simulated to determine their performance across a variety of realistic scenarios for how data may accumulate during the trial and designed to offer similar power with the expected 7% treatment effect. The alternative design was published along with a prospectively defined plan to collect the data that would have been necessary to conduct its analysis plan.^9^ Therefore, during the conduct of the SHINE trial, data snapshots were frozen whenever an interim analysis for the alternative Bayesian design would have occurred. This has now allowed the alternative Bayesian design to be virtually executed, using the actual trial data, exactly as would have happened had the Bayesian design been selected for implementation.

One of the barriers to the uptake of Bayesian adaptive designs for confirmatory trials is relative lack of experience with how they may perform and compare, in practice, to a frequentist design. The SHINE trial is a rare opportunity to compare more commonly used trial features (blinded sample size re-estimation and boundary based rules for efficacy/futility stopping) to a counterfactual Bayesian design, both prospectively designed by the same lead investigators around the same primary scientific question and goals. There have been only a few published examples of randomized clinical trials reconsidered, in a retrospective manner, with alternative trial designs and we are unaware of any reports based on a predefined alternative.^10–12^ Here, we report the results from the virtually executed Bayesian alternative design of the SHINE trial.

## Methods

The SHINE trial was a randomized clinical trial investigating intensive glucose control versus standard of care for stroke patients with hyperglycemia.^13^ The primary efficacy endpoint was a dichotomous assessment of a favorable neurologic outcome at 90 days. A favorable outcome for a patient was defined based on a sliding dichotomy of the modified Rankin scale (mRS), dependent on baseline stroke severity.^14,15^ The maximum sample size was 1400 patients, which provided 80% power to detect a clinically meaningful improvement in the proportion of patients achieving favorable outcomes from 25% on the standard of care arm to 32% for the intensive glucose control arm based on a two-sided frequentist test at the 5% level.

### SHINE Trial

The SHINE trial design that was executed was a group sequential trial with two-sided efficacy and futility stopping boundaries along with response-adaptive randomization. The stopping boundaries were defined using the gamma family spending functions and closely resembled an O’Brien-Fleming boundary.^16^ Futility boundaries were considered non-binding, meaning that if a futility boundary was crossed, the trial could be continued without inflation of the Type 1 error rate. Interim analyses were planned when 500, 700, 900, and 1,100 consecutively randomized patients had reached the 90-day follow up period. Multiple imputation was used to address missing primary outcome data for all interim and final analyses. A blinded sample size re-estimation was planned to occur prior to the first interim analysis. The blinded sample size re-estimation could increase the trial’s maximum sample size if the proportion of patients experiencing a favorable outcome was higher than expected. The full design is specified in the protocol and the statistical analysis plan which are available as supplements to the main trial publication.^17^

### SHINE Alternative Design

The alternative design was a Bayesian adaptive “Goldilocks” trial design.^18^ The sample size re-estimation and early stopping included in the frequentist SHINE trial are both focused on the goal of “right-sizing” the trial in response to the observed trial data. Similarly, the Goldilocks design is a sample size selection approach that aims to “right-size” the number of patients randomized between a prespecified minimum and maximum in response to the observed trial data. This design included more frequent interim analyses with the first interim analysis scheduled to occur after 500 patients randomized and subsequent interims planned after every additional 100 patients randomized. Trials using this approach have been used in the support of multiple regulatory approvals by the US Food and Drug Administration.^19–23^

With the interim analyses timed according to the number of patients randomized, not all patients included in the interim analyses would have known 90-day primary endpoint outcomes. However, patients in the trial were also being assessed for a favorable neurologic outcome at 6 weeks, and so the Bayesian design made use of that earlier information with a longitudinal model linking the 6-week and the 90-day outcome. For patients included in the interim analysis who have an unknown 90-day outcome but a known 6-week outcome, the longitudinal modeling is used to impute their 90-day outcome. The longitudinal model is based on the currently observed data and takes into account uncertainty in how the 6-week outcome predicts the 90-day outcome. Patients without a 6-week or 90 day outcome contribute no outcome information at the interim analyses.

At each interim analysis, two predictive probabilities of trial success are calculated.^24^ The first is the predictive probability of trial success with the currently randomized patients. If this predictive probability is high, accrual may stop accrual for early success. In this case, all randomized patients would complete follow-up for their primary endpoint, and the final analysis is then conducted on that complete dataset. The rationale is that if the currently randomized sample size provides sufficient power for the treatment effect being observed in the trial then additional trial resources are not required. The second predictive probability is the predictive probability of trial success at the maximum sample size of 1400 patients. If this value is low, the trial may stop early for futility. The rationale in this case is that the maximum amount of trial resources would still not provide an adequate chance of trial success and as such, the trial should end based on an inability to successfully address its primary objective. For the SHINE trial, the predictive probability at the current sample required to stop accrual early for success was set at 99% and the predictive probability at the maximum sample size required to stop the trial early for futility was set at 5%. The final analysis was a Bayesian posterior probability of superiority. In order to account for the multiple interim analyses, the final critical value required for statistical significance was set at a Bayesian posterior probability of superiority of at least 97.9%. This value was selected to control the overall one-sided Type I error of the trial to less than 2.5%. Thus these futility rules are a binding futility stop. This design and its operating characteristics are completely described elsewhere.^9^

### Analysis Methods

This is a pre-planned, secondary analysis of the SHINE trial. The statistical and data management team for the SHINE trial preserved snap-shots of the accumulating enrollment and outcome data consistent with the timing of interim analyses specified in the alternative Bayesian design. After the SHINE trial was complete, these datasets were used to virtually conduct the interim analyses prespecified in the alternative design. All computations for the alternative design were performed in R. The code is available at http://hdl.handle.net/2027.42/167625.

## Results

### SHINE trial conduct as enrolled

The blinded sample size re-estimation did not result in an increase to the study’s sample size and the study proceeded with the planned interim analyses. A summary of the observed interim analysis results along with the corresponding boundaries for each of the SHINE trial looks is shown in Table 1. The SHINE trial crossed its pre-specified futility boundary at its third interim analysis with 936 patients randomized and 900 consecutively randomized patients completing the 90-day follow up period. The adjusted estimate of the treatment effect favored intensive glucose control by 0.1%. However, the Data Safety and Monitoring Board (DSMB) did not recommend that the trial stop at this interim and the trial continued. The pre-specified futility boundary was crossed again at the fourth interim analysis with 1137 patients randomized and 1100 consecutive patients included in the analysis. At this interim, the DMC recommended that the trial stop for futility. The final total sample size of SHINE was 1151 patients. The final primary analysis result showed that 21.6% (123/570) of patients in the control arm and 20.5% (119/581) of patients in the treatment arm had a favorable 90-day outcome.

**Table 1:** SHINE frequentist interim analyses and boundaries

| N<br>Enrolled | N<br>Analyzed | Stopping Boundaries |  | N with 90<br>day<br>outcome<br>in analysis<br>population | Favorable outcome<br>(Unadjusted) |  | Adjusted<br>Risk<br>Difference*<br>(p value) |
| --- | --- | --- | --- | --- | --- | --- | --- |
|  |  | Efficacy<br>Z value<br>(p value) | Futility<br>Z value<br>(p value) |  | Intensive | Standard |  |
| 579 | 500 | 2.97<br>(0.003) | 0.06<br>(0.949) | 482 | 68/254<br>(26.8%) | 53/228<br>(23.3%) | 2.8%<br>(0.134) |
| 771 | 700 | 2.86<br>(0.004) | 0.13<br>(0.896) | 677 | 83/344<br>(24.1%) | 78/333<br>(23.4%) | 0.9%<br>(0.557) |
| 936 | 900 | 2.65<br>(0.008) | 0.45<br>(0.652) | 869 | 97/444<br>(21.9%) | 97/425<br>(22.8%) | 0.1%<br>(0.957) |
| 1137 | 1100 | 2.42<br>(0.016) | 1.05<br>(0.293) | 1061 | 115/540<br>(21.3%) | 115/521<br>(22.1%) | -0.1%<br>(0.915) |

\*with multiple imputation for missing outcome data

### SHINE trial conduct under alternative design

While the SHINE trial crossed its futility boundary with 936 patients enrolled, the Bayesian alternative design crossed the futility boundary at its 4th interim analysis with 800 patients randomized and 715 patients complete for the primary endpoint. A summary of the results of each of the Bayesian alternative design interim analyses is provided in Table 2.

**Table 2:** Bayesian Alternative Design Interim Analyses

|  |  | Good Outcome Proportion |  | Predictive probability of Trial Success |  |
| --- | --- | --- | --- | --- | --- |
| N enrolled | N with 90 day outcome | Treatment | Control | Current Sample Size (Success)<br>Success Stopping Bound: > 99% | Maximum Sample Size (Futility)<br>Futility Stopping Bound: < 5% |
| 498 | 432 | 26.6% | 24.6% | <1% | 21.1% |
| 579 | 515 | 25.7% | 22.3% | <1% | 39.7% |
| 700 | 621 | 25.1% | 23.4% | <1% | 15.6% |
| 800 | 715 | 23.1% | 22.80% | <1% | 2.0% |

Figure 1 shows the predictive distribution of the proportion of patients experiencing a favorable outcome with the currently randomized patients as well as at the maximum sample size of 1400 patients. Considering the similarity of the observed 90-day outcomes the predictive probability of success at the current sample size was essentially 0% and the predictive probability of success at the maximum sample size was 2%. This latter quantity was less than 5% required to stop the trial for futility. Results were not sensitive to the longitudinal model that imputed 90-day outcomes for patients only observed to be complete through 90-days. Repeating the alternative design execution without the longitudinal model resulted in the trial stopping for futility at the same interim analysis.

**Figure 1:**
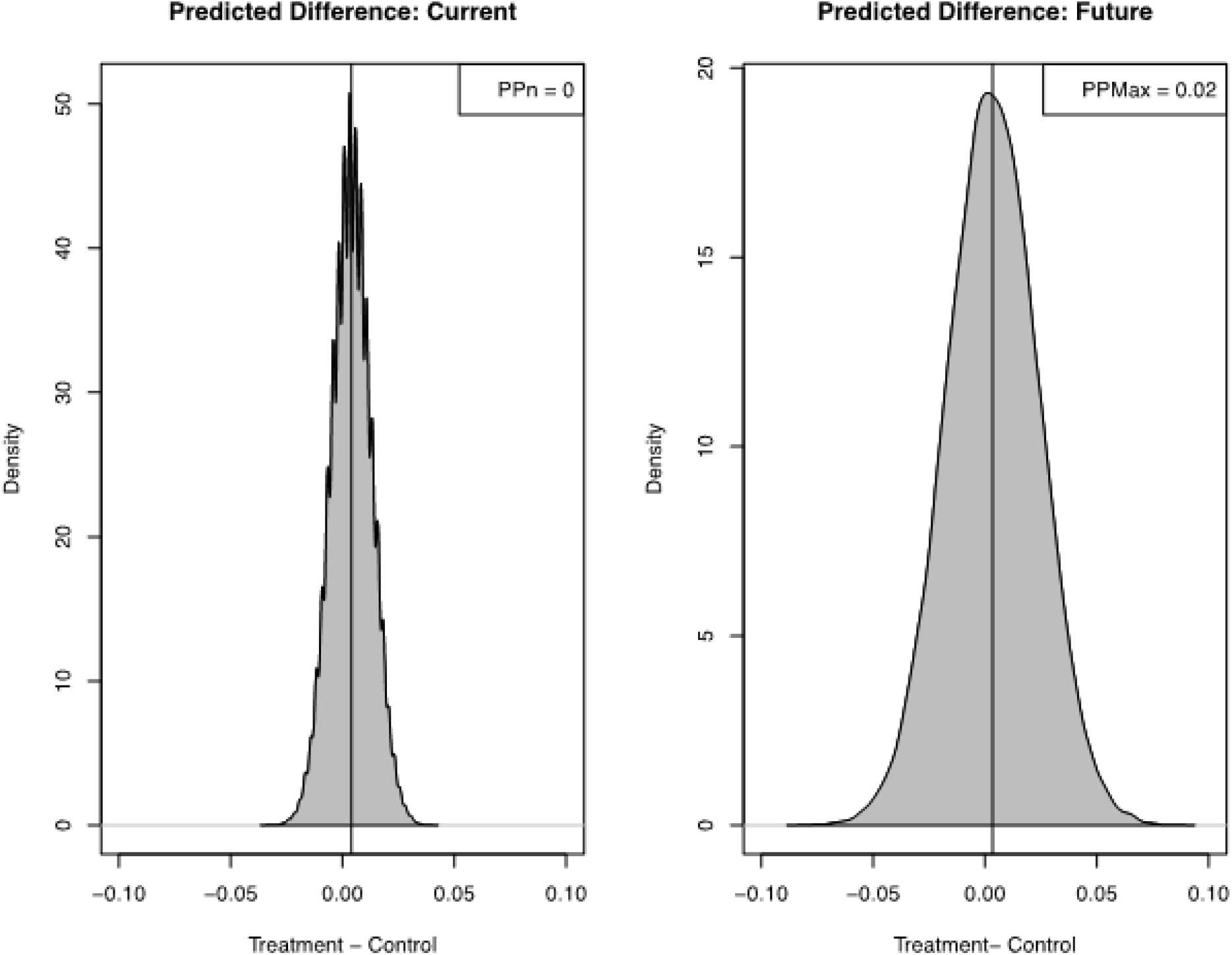
Posterior Density Plots. Density plots for the posterior probability of superiority of treatment at current sample size (PPn) and at the design based maximum sample size (PPMax).

Table 3 shows the final results of the primary and key secondary outcomes at the time each trial crossed their futility boundary, not at the time each trial actually stopped enrollment as the DMC decision could not be recreated for the alternative design. Estimates do vary slightly, but overall inference is similar between the two trial designs across all outcomes.

**Table 3:** Trial Outcomes Based on Design Based Stopping

|  | **SHINE frequentist**<br>**N=936** |  | **SHADOW SHINE**<br>**N=800** |  |
| --- | --- | --- | --- | --- |
|  | **Intensive**<br>**N=483** | **Standard**<br>**N=453** | **Intensive**<br>**N=408** | **Standard**<br>**N=392** |
| Primary Efficacy Outcome N(%) |  |  |  |  |
| Favorable* | 102 (21.1) | 102 (22.5) | 89 (21.8) | 89 (22.7) |
| Missing** | 18 (3.7) | 12 (2.7) | 14 (3.4) | 11 (2.8) |
| Unadjusted Risk Difference<br>% (95% CI) | -1.4 (-6.7, 3.9) |  | -0.9 (-6.7, 4.9) |  |
| Adjusted*** Risk Difference %(95%<br>CI) | -0.05 (-2.6 , 2.5) |  | 0.3 (-2.5, 3.0) |  |
| P - value for adjusted<br>analysis | 0.97 |  | 0.86 |  |
| Secondary Efficacy Outcome n/N(%) |  |  |  |  |
| Favorable NIHSS (0 or 1) | 125/293<br>(42.7) | 144/306<br>(47.1) | 109/247<br>(44.1) | 131/270<br>(48.5) |
| Unadjusted Risk Difference<br>% (95% CI) | -4.4 (-12.4, 3.6) |  | -4.4 (-13.0, 4.2) |  |
| Favorable Barthel Index (95-100) | 222/408<br>(54.4) | 218/387<br>(56.3) | 193/344<br>(56.1) | 194/336<br>(57.7) |
| Unadjusted Risk Difference- % (95%<br>CI) | -1.9 (-8.8, 5.0) |  | -1.6 (-9.1, 5.8) |  |
| SSQOL N | 366 | 353 | 312 | 304 |
| Median (IQ) | 3.73<br>(2.86, 4.39) | 3.70<br>(3.07, 4.46) | 3.71<br>(2.84, 4.39) | 3.73<br>(3.06, 4.50) |
| Difference in Medians- (95% CI) | 0.03 (-0.18, 0.23) |  | -0.02 (-0.23, 0.23) |  |
| Primary Safety Outcomes - N (%) |  |  |  |  |
| Death | 44 (9.1) | 49(10.8) | 37 (9.1) | 41 (10.5) |
| Risk Difference- % (95% CI) | -1.7 (-5.6, 2.1) |  | -1.4 (-5.5, 2.7) |  |
| Severe Hypoglycemia<br>($<40\text{mg/dL}$) | 12 (2.5) | 0 | 10 (2.5) | 0 |
| Risk Difference -% (95% CI) | 2.5 (1.1, 3.9) |  | 2.5 (1.0, 4.0) |  |

## Discussion

Both trial designs reached the same conclusion of futility, while the Bayesian alternative design reached that conclusion earlier in the course of the participant accrual.^17^ Operating characteristics estimated prior to trial conduct using numerical simulation under the null hypothesis of no treatment benefit showed that the SHINE trial had an expected sample size of 1039 and that the Bayesian alternative had an expected sample size of approximately 750.^9^ The implemented, frequentist, SHINE design first crossed its futility boundary with 936 patients randomized and the alternative design crossed its futility boundary with 800 patients randomized. The difference is driven by the enrollment rate and the implemented SHINE design needing to wait until the subjects reached the 90-day follow up. Given the data observed during the actual trial, it was more likely than not that both trial designs would stop for futility. Under the null, the SHINE trial had a 73% probability of early futility and the alternative design had an 91% probability of early futility.

One aspect of the trial execution not reflected with this virtual execution of the alternative design is the DSMB review of the interim analyses and recommendations to continue accrual or stop for futility. In this trial, the stakeholders were interested in providing a definitive answer to the acute stroke community. As such, there was a strong desire to ensure residual uncertainty was minimized. Examples include the possibility that some acute stroke patients (mild or severe) might have possible benefit from the intensive glucose management strategy on subgroup analyses. One may speculate that a DSMB would have the same concerns with the Bayesian design, particularly if it suggested stopping even earlier in the course of the trial.

Concerns regarding Bayesian adaptive design in confirmatory phase trials include the operational complexity of the frequent interim analyses, uncertainty with how the Bayesian algorithms may perform given the observed data, and the scientific validity of the decision making. While some investigators equate Bayesian models with informative priors, in reality the goal of this method was to fit longitudinal models in a way that was driven by the goal of predicting trial success and failure. It is beyond the scope of the current investigation to discuss whether available frequentist models could be used for this form of within trial decision making, it is notable that they usually are not. While traditionally only participants with final outcomes contribute to frequentist interim analyses, the partial information from early outcomes was used to inform the Bayesian interim analysis. These outcomes can be potentially informative as long as the uncertainty within it is properly accounted for and understood.

This example shows that it was feasible to collect and freeze the data necessary to conduct the more frequent interim analyses. While not carried out in this virtual execution, the reporting to the DSMB of the interim analysis results can usually be accomplished in a matter of several business days after the interim milestones are achieved with proper systems in place. This example also illustrates throughout the interim analyses how the Bayesian algorithm reacted to both the observed data and the uncertainty in the data yet to be collected and resulted in the same scientific conclusion as the group sequential SHINE trial. This conclusion was reached more efficiently, in terms of sample size, driven by the more frequent interim analyses and therefore greater opportunity for early stopping. There are multiple examples during COVID-19 showing that rapid, high quality interim analyses can be conducted in prespecified Bayesian adaptive trials.^25^

The group sequential design and the Bayesian Goldilocks sample size algorithm are similar in spirit. Both techniques aim to limit patient resources in the case of overwhelming efficacy or in the case of futility in the observed data. It is expected that this head to head comparison of the two approaches resulted in the same trial conclusion, especially considering that the observed trial data did not show a difference between the treatment strategies. As familiarity with the design and operationalization of Bayesian adaptive designs increases we hope to see more innovative features incorporated into confirmatory trials.

This work has important limitations. First, this was a shadow trial conducted post hoc to see how it would have turned out,that used actual data that would have been available at each interim analysis. In practice, however sufficient resources do not exist to run multiple, concurrent clinical trials to assess differences in designs. Even if that had occurred here, the common practice of DSMBs would have likely prompted the cessation of the parallel trial. Second, the trial studied was relatively simple -a two group comparison with a binary primary endpoint. Similar prospective comparisons of more complicated designs such as those studying multiple doses or having multiple stages would be more challenging and likely could not utilize the exact sequence of enrollments and timings of outcomes. Third, while ending a trial early for futility when results are unambiguous is beneficial for the patients, clinical community, and can reduce the financial cost, the current method of submitting, reviewing, and issuing grants to academics is not as well suited to this; whereas the value proposition for a pharmaceutical company bearing all of the investment is clearer.^26^ Finally, this study was unique amongst publicly funded U.S. clinical trials in that it had substantial time and monetary resources dedicated to trial design and simulation through the ADAPT-IT project. While not a limitation of this study per se, it represents a limitation of the current paradigm for proposing, planning and designing NIH-funded clinical trials in the United States.

In conclusion, the two possible SHINE designs performed similarly when exposed to the actual patient responses observed in the trial. The Bayesian design reached a conclusion of futility earlier, consistent with the operating characteristics of the null scenario simulated prior to starting the trial and in part due to the Bayesian interims also incorporating incomplete data through a longitudinal model. Trialists should consider developing, simulating, publishing and assessing counterfactual designs in parallel with actual study conduct to gain additional knowledge and expand understanding of more innovative randomized trials.

## Data Availability

Dr. Durkalski and Ms. Broglio can attest to the integrity of the data analysis and access. While the investigators used the trial database, other individuals can obtain the trial data from the National Institutes of Neurological Disorders and Stroke Archived Clinical Research Datasets available online. The code to run the simulations is available at the DeepBlue University of Michigan archive.

https://www.ninds.nih.gov/Current-Research/Research-Funded-NINDS/Clinical-Research/Archived-Clinical-Research-Datasets

http://hdl.handle.net/2027.42/167625

## Acknowledgments

None.

## Ethics

The research was conducted with a dataset free of personal identifiers. The data coordinating center did not have access to trial participant identities and the investigators could not link the data to individuals. This was considered a non-regulated activity by the University of Michigan IRBMED.

## Data Access and Responsibility

Dr. Durkalski and Ms. Broglio can attest to the integrity of the data analysis and access. While the investigators used the trial database, other individuals can obtain the trial data from the National Institutes of Neurological Disorders and Stroke Archived Clinical Research Datasets available online

## Authors’ contributions

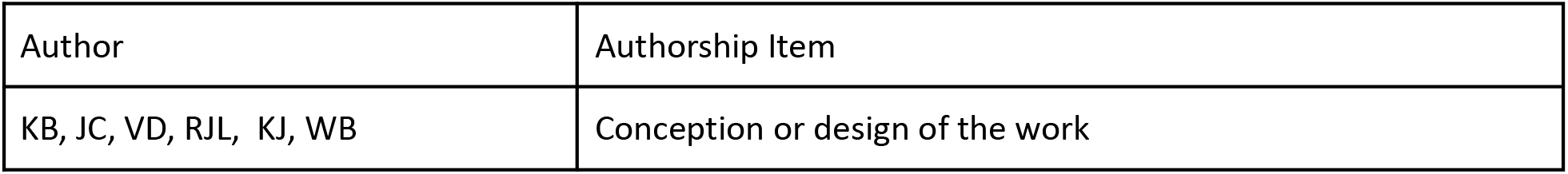
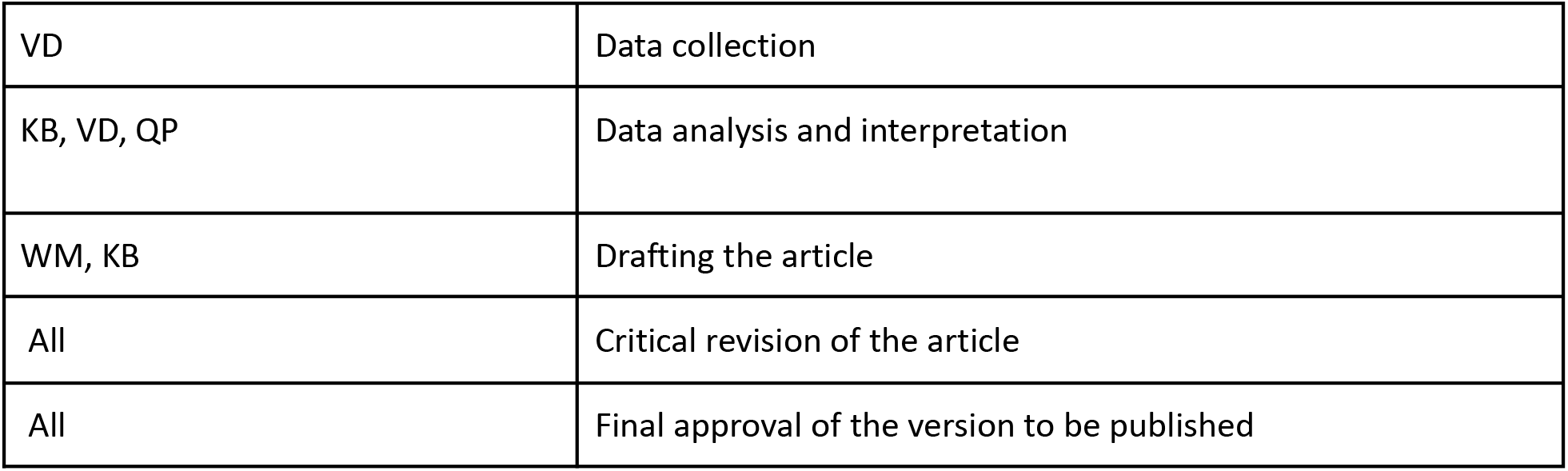

| Author | Authorship Item |
| --- | --- |
| KB, JC, VD, RJL, KJ, WB | Conception or design of the work |
| VD | Data collection |
| KB, VD, QP | Data analysis and interpretation |
| WM, KB | Drafting the article |
| All | Critical revision of the article |
| All | Final approval of the version to be published |

